# Association of sexual orientation outness and recent homophobic violence with not being on antiretroviral treatment: Analysis of a Latin American Survey in men who have sex with men living with HIV

**DOI:** 10.64898/2026.04.22.26351515

**Authors:** Juan C. Enciso-Durand, Alfonso A. Silva-Santisteban, Michael Reyes-Díaz, Luis Huicho, Carlos F. Cáceres

## Abstract

**Objectives:** In Latin America, up-to-date information to monitor UNAIDS 95-95-95 HIV targets in key populations, such as men who have sex with men, is limited. Elsewhere, structural homophobia restricts access to ART. Conceptual frameworks suggest that intersecting forms of violence and discrimination may negatively influence HIV care outcomes through psychosocial and structural pathways, although empirical evidence remains limited. The study aimed to assess whether sexual orientation outness and recent homophobic violence are associated with not being on ART among Latin American MSM living with HIV.

**Methods:** This cross-sectional study is a secondary analysis of data from LAMIS-2018, including 7,609 MSM aged 18+ with an HIV diagnosis ≥1 year prior from 18 Latin American countries. Participants self-reported ART status, sociodemographic characteristics, homophobic violence, and sexual orientation outness. Bivariate and multivariate logistic regressions identified those factors associated with not being on ART.

**Results:** Nine percent of MSM with HIV were not on ART, 18% reported low sexual orientation outness, and 27% experienced homophobic violence, especially in Andean and Central American countries. Not being on ART was associated with recent homophobic violence (aPR=1.25), low outness (aPR=1.22), unemployment (aPR=1.27), and residence in the Andean subregion (aPR=1.87), Mexico (aPR=1.28), or the Southern Cone (aPR=1.45) versus Brazil. Protective factors included being older (25–39: aPR=0.72; >39: aPR=0.49), living in large cities (aPR=0.72), having a stable partner (aPR=0.78), and university education (aPR=0.74).

**Conclusions:** Recent homophobic violence and low sexual orientation outness were associated with not being on ART among MSM in Latin America. While access varies across countries, structural factors such as stigma and violence may limit engagement in care. Addressing these barriers alongside strengthening health systems may be key to improving ART uptake and advancing progress toward the 95–95–95 targets.

## Introduction

The HIV pandemic remains a public health issue in Latin America (LA), one of the few regions where HIV incidence has not decreased[1]. By 2024, the Joint United Nations Program on HIV/AIDS (UNAIDS) reported 39.9 million people living with HIV (PLHIV) globally[2]. In 2021, 2.2 million PLHIV were estimated in the region[3], 69% of whom were thought to be on antiretroviral therapy (ART). Key populations such as transgender women and men who have sex with men (MSM) are the most affected by HIV, with regional prevalence rates exceeding 13%[4] and ART coverage rates lower than overall figures[4]. Notably, since 2020, LA has experienced a 9% increase in new HIV cases among these groups—an upward trend not seen in most other regions worldwide[5]. Furthermore, discriminatory attitudes persist toward PLHIV and key populations[6], negatively impacting public health[7] and efforts to fight HIV[8].

UNAIDS set the global 95-95-95 targets to fight HIV by 2030, aiming for 95% of people living with HIV to be diagnosed, 95% of them to receive antiretroviral treatment, and 95% of those on treatment to achieve viral suppression[9]. The LA region is far from meeting those targets[10], although appropriate monitoring is difficult given limitations in the availability of strategic information on MSM and TGW[6], particularly on the loss to follow-up from ART(i.e. dropout or discontinuation)[11] or on “non-linkage” to ART[12,13]. The 95-95-95 targets may not be achieved if the high dropout rates persist[14–16]. The most recent data on ART loss to follow-up in LA, estimating a range from 6% to 36%, pertain to the period 2008–2012 for Brazil, Perú, Central America and the Caribbean[17].

Access to HIV prevention and treatment services largely depends on the level of development and capacity of national health systems, as stronger systems are better able to ensure availability, continuity of care, and equitable access[18]. In addition, the literature indicates that homophobic violence, as well as social and mental health factors, play a critical role in shaping access to HIV testing[19] and ART[20]. For instance, substance use has been shown to negatively affect individuals’ ability to remain on ART[21–23]. Similarly, mental health conditions[24,25] and the absence of social support are associated with a higher likelihood of treatment discontinuation[26,27]. Some individuals may interrupt treatment to avoid being identified as PLHIV when accessing services[13,28], while fear of discrimination following disclosure of serostatus to family, friends, or partners can further contribute to disengagement from care[28,29]. Additionally, experiences of discrimination within healthcare settings can erode trust in services, thereby undermining treatment continuity[26,28,30,31]. Taken together, these findings suggest that, beyond health system limitations, mental health problems, discrimination, and violence are central to understanding both access to HIV services and continuity in antiretroviral treatment.

Conceptual frameworks, such as the model proposed by Quinn et al.[32], suggest that different and overlapping forms of violence, including childhood sexual abuse, intimate partner violence, and structural or community-level violence—may interact to produce compounded effects on HIV-related outcomes, such as engagement in care, adherence to ART, loss to follow, and viral suppression (Figure 1). These influences may operate through several mechanisms, including deteriorated mental health, internalized stigma, symptoms of post-traumatic stress, and diminished trust in health systems. Nevertheless, although these pathways are increasingly acknowledged in theoretical discussions, empirical evidence supporting them remains scarce, particularly in LA.

**Figure 1.**
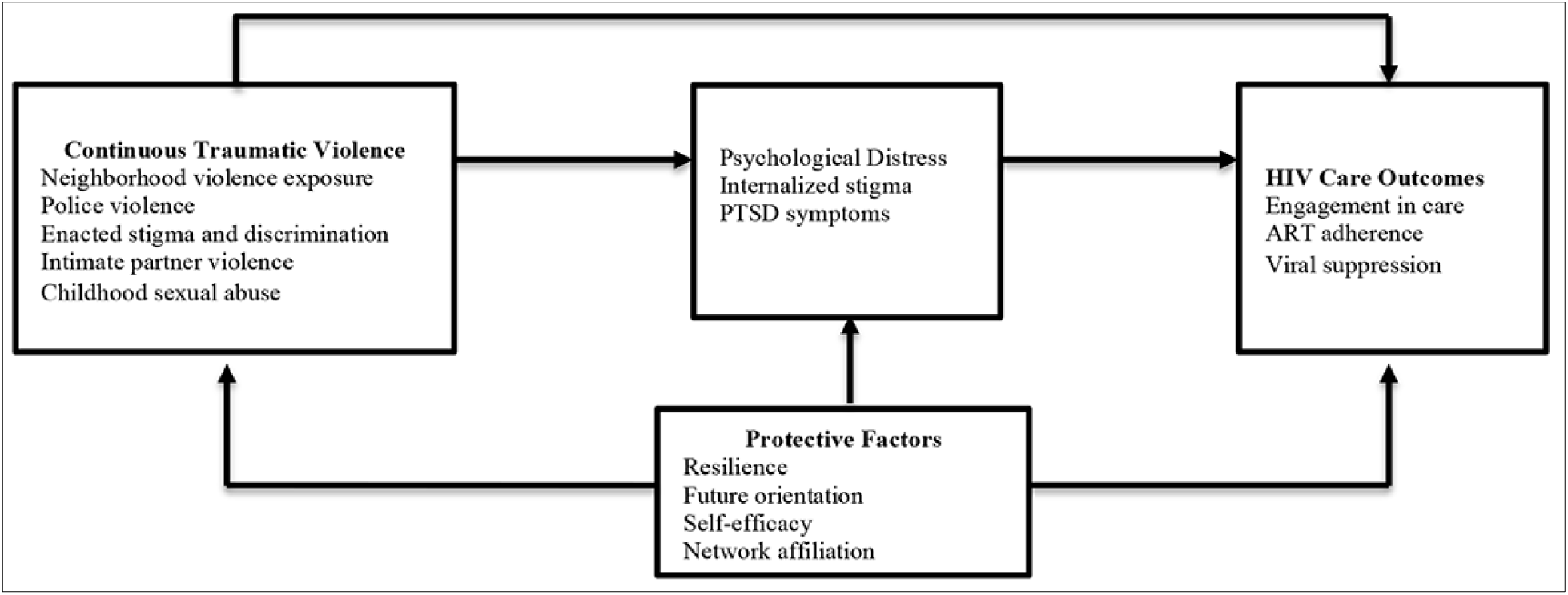
Conceptual framework linking violence exposure, psychological distress and HIV care outcomes among MSM (Quinn et al., 2021).

Studies on mental health, violence and discrimination in LA in recent years are scarce compared to those in other regions such as Sub-Saharan Africa. The study by Boni et al., which examines the issue in several LA countries[23], found a dropout rate of 6%, which increased to 8% and 12% among alcohol (OR=1.2) and drug (OR=1.3) users, respectively. Another regional study from 2015 found an 18% dropout rate but did not assess the association with mental health or discrimination indicators[33]. Up to 17% dropout has been reported in Colombia and Brazil, with limited evaluation of potentially associated mental health and discrimination factors[22,34]. A study conducted in Mexico showed that experiencing homophobic violence decreases adherence to ART[35]. This situation reaffirms the UNAIDS’ conclusion on the need for more strategic and up-to-date information on the problem in LA[6].

The “Latin America MSM Internet Survey” (LAMIS-2018) was the first online survey providing regional information on HIV prevention, access to ART, sexual health, stigma and discrimination, and mental health among MSM[36]. The survey was administered simultaneously across 18 LA countries in 2018. Given the limited strategic information on MSM living with HIV in the region, the data provided by LAMIS-2018 allows up-to-date insights into ART loss to follow up. The aim of this study is to assess whether recent homophobic violence and sexual orientation outness, as expression of current exposure to structural homophobia, are associated with not being on ART among MSM, either due to not initiating treatment or to discontinuing it in LA.

## Methods

### Study design and Setting

This is a cross-sectional, observational study based on a secondary analysis of data. The LAMIS-2018 survey was implemented by researchers from the “Ibero-American Network of Studies on Gay Men, Other Men Who Have Sex with Men, and Transgender People” (RIGHT PLUS)[36,37]. The survey was conducted online from January to May 2018 in Argentina, Bolivia, Brazil, Chile, Colombia, Costa Rica, Ecuador, El Salvador, Guatemala, Honduras, Mexico, Nicaragua, Panama, Paraguay, Peru, Surinam, Uruguay, and Venezuela. Each country had a local team of researchers leading the process. The survey was available in Spanish, Portuguese, and Dutch, and a non-probabilistic quota sampling method was used. The survey was disseminated through dating apps for men, websites, social media, clinics, non-governmental organizations, and leisure spaces frequented by MSM. After removing inconsistent or invalid data, LAMIS included 64,655 valid cases. Further details on the study methodology are available in the published article[38], and additional results can be found in the full report[36].

### Participants

Selection criteria for the regional study included: identifying as a male or transgender male, being sexually attracted to men, and/or having ever had sexual interactions with other men; residing in the countries of interest; and being over 18 years old. For the current study, an additional criterion was having been diagnosed with HIV at least 1 year before data collection (individuals diagnosed within the past year were excluded to avoid bias related to recent detection or bureaucratic treatment delays). These exclusions occurred in similar proportions across all subregions. A total of 7,609 observations met the inclusion criteria. The process of participant selection is shown in Figure 2.

**Figure 2.**
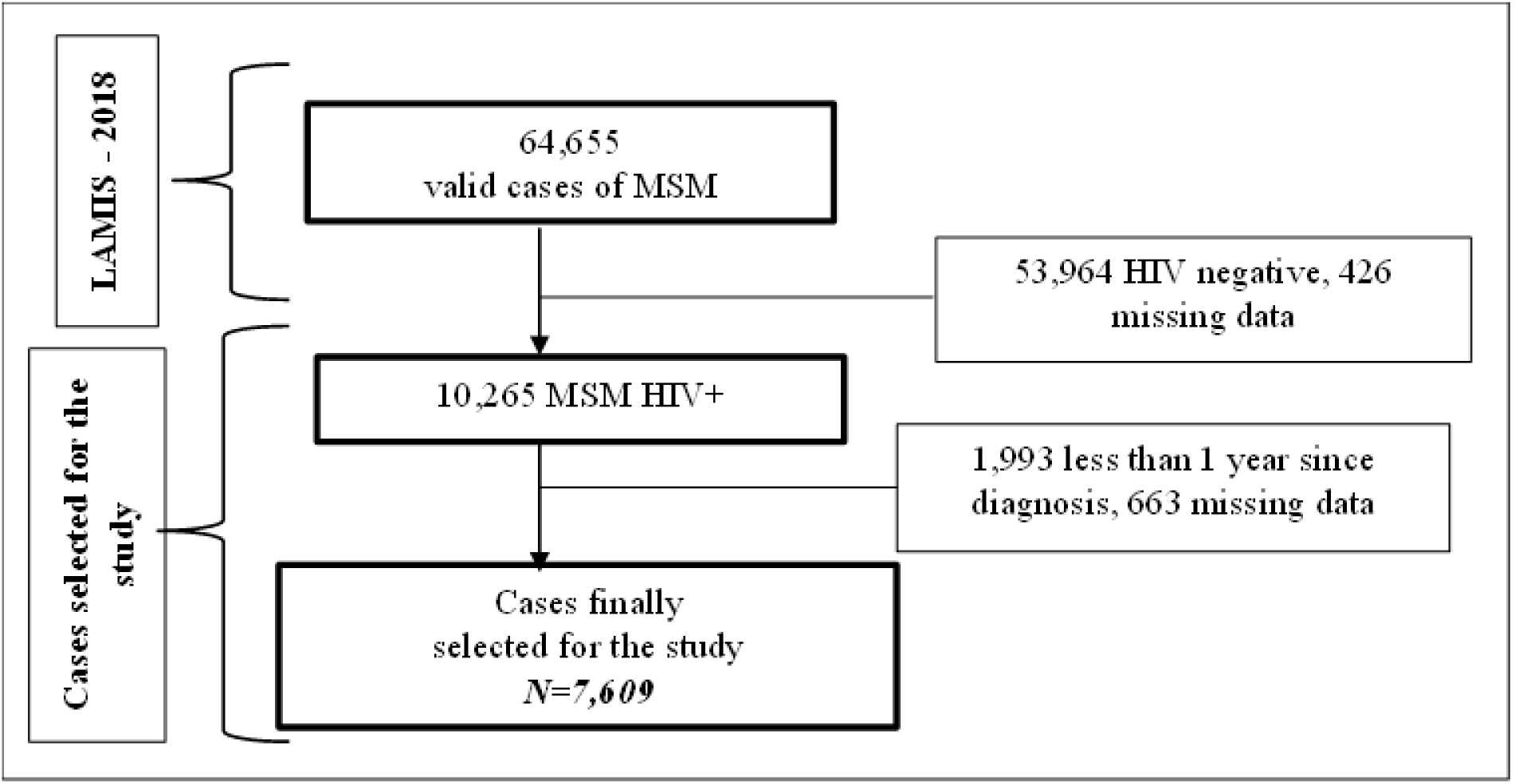
Study flowchart.

### Variables and data source

The outcome variable in our study was “ART status,” with two possible statuses: “on ART” or “not on ART”. This was determined based on the responses to the following questions: “Did you start treatment?” and “Are you currently on treatment?”. The status was defined as “on ART” when participants responded that they had started and were currently under treatment. Any other combination of responses was considered as “not on ART”.

The independent variables analyzed were “Sexual orientation outness**”** and “Recent homophobic violence.” Outness was operationalized as the proportion of individuals in the participant’s social environment who were aware of his attraction to men[39], as assessed by the question: ‘Thinking about the people who know you, what proportion of them knows that you are attracted to men?”. Three response categories were considered: “All or almost all know” “Some know”, and “Few or none know”[36]. The second variable, “Homophobic violence” (called homonegative experiences in some LAMIS publications[36,40]) refers to verbal abuse, threats, or physical violence suffered due to sexual orientation, in public or private spaces[35] and is a proxy for exposure to structural homophobic violence. Responses were dichotomized to identify experiences of recent verbal abuse, threats, or physical violence if the participant reported experiencing physical, verbal abuse, or threats in the past 4 weeks; otherwise, if the events occurred more than 4 weeks ago or never, they were considered as ‘absence of recent violence’. We focused on recent violence for two reasons: it is considered as a proxy to current exposure to structural homophobic violence[41]; and longer recall periods, such as 12 months, are more susceptible to recall bias due to memory decay, telescoping, and under-reporting[42].

Potential confounding variables, based on available scientific evidence[24,30,31] were included. The first was symptoms of anxiety and depression, assessed using the PHQ-4[43] and dichotomized into Normal-Low (<6 score) and Moderate-Severe (≥6 score). Alcohol dependence was measured using the CAGE-4 questionnaire[44], classified as No Dependence (<2 score) and Dependence (≥2 score). In this study, Cronbach’s alpha values for the PHQ-4 and CAGE-4 were 0.89 and 0.60, respectively. Recent drug use was also analyzed, including substances such as synthetic cannabinoids, ecstasy, amphetamines, heroin, cocaine, and others. Recent use was defined as consumption reported within the past 24 hours or 7 days, while no recent use referred to never having used or use occurring more than 7 days prior. Additionally, sociodemographic variables were included: age, country, city size, stable partner, education level, and employment status. Countries were grouped into subregions following the regional classification used in the main LAMIS-2018[36] analysis: Brazil, Andean Countries (Bolivia, Ecuador, Peru, Colombia and Venezuela), Mexico, Southern Cone (Argentina, Chile, Paraguay, and Uruguay) and Central America and the Caribbean (Costa Rica, El Salvador, Guatemala, Honduras, Nicaragua, and Panama and Suriname).

### Statistical Power

Statistical power was calculated using the group proportion comparison module of Epidat, version 4.2. For “Recent homophobic violence” a power of 99% was estimated, using a “Not on ART” proportion of 8.3% in the unexposed group and 11.6% in the exposed group, with a group ratio of 2.7. For “Sexual orientation outness “ using the “Few or Non know” category as the exposed group, a power of 76% was obtained, with an event proportion of 8.7% in the unexposed group and 11.1% in the exposed group, with a group ratio of 4.4. The calculated figures were close to or above the recommended 80% threshold[45], ensuring that the study had sufficient statistical power to avoid Type II error.

### Data management and statistical analysis

The RIGHT PLUS technical committee provided the dataset along with user instructions. Analyses were conducted using STATA 18. A descriptive analysis of the variables of interest was performed using frequency and proportion analyses. Associations between study variables were initially explored using the Chi-square (χ²) test. Adjusted and unadjusted prevalence ratios (PRs) were estimated using generalized linear regression models with a logarithmic link function, and robust standard errors were applied to account for heteroscedasticity. Models were adjusted for demographic variables and potential confounders, including alcohol use, drug use, and symptoms of anxiety and depression. Given the large sample size, results were interpreted considering not only statistical significance but also the magnitude and precision of effect estimates, to identify findings of practical relevance.

### Ethical considerations

Ethical approval for the regional study was granted by five local ethics committees across the 18 countries where the survey was conducted, along with verbal informed consent from participants[38]. For this secondary data analysis, the anonymized database was requested through the regional study website of LAMIS-2018 (https://www.coalitionplus.org/lamis/). Approval was obtained, and access to the dataset was granted on October 2, 2023. The authors did not have access to information that could identify individual participants. Ethical approval for this study was obtained from the Institutional Ethics Committee of the Universidad Peruana Cayetano Heredia (approval code: CIEI-196-20-24) on May 7, 2024. This study did not generate new data; it exclusively used previously collected and available data for the analysis. As the dataset was fully anonymized, no informed consent was required for this secondary analysis.

## Results

### Participants overview

Out of 7,609 participants, 65% were between 25 and 39 years old, a quarter over 39 years old, and less than 10% under 25 years old. By subregion, 29% were from Brazil, 23% from the Andean countries, 22% from Mexico, 21% from the Southern Cone countries, and 5% from Central America and the Caribbean. The majority reside in large cities with more than 100,000 people (89%). Nearly 70% were single and had a university education or higher. Around 80% were employed. Regarding the study variables, 9.2% were not on ART and 27% reported experiencing homophobic violence in the last 4 weeks. In terms of sexual orientation outness, 53% reported that “All or almost all know” about their orientation, 29% said “Some know” and 18% reported that “Few or none know” (Table 1).

**Table 1.**
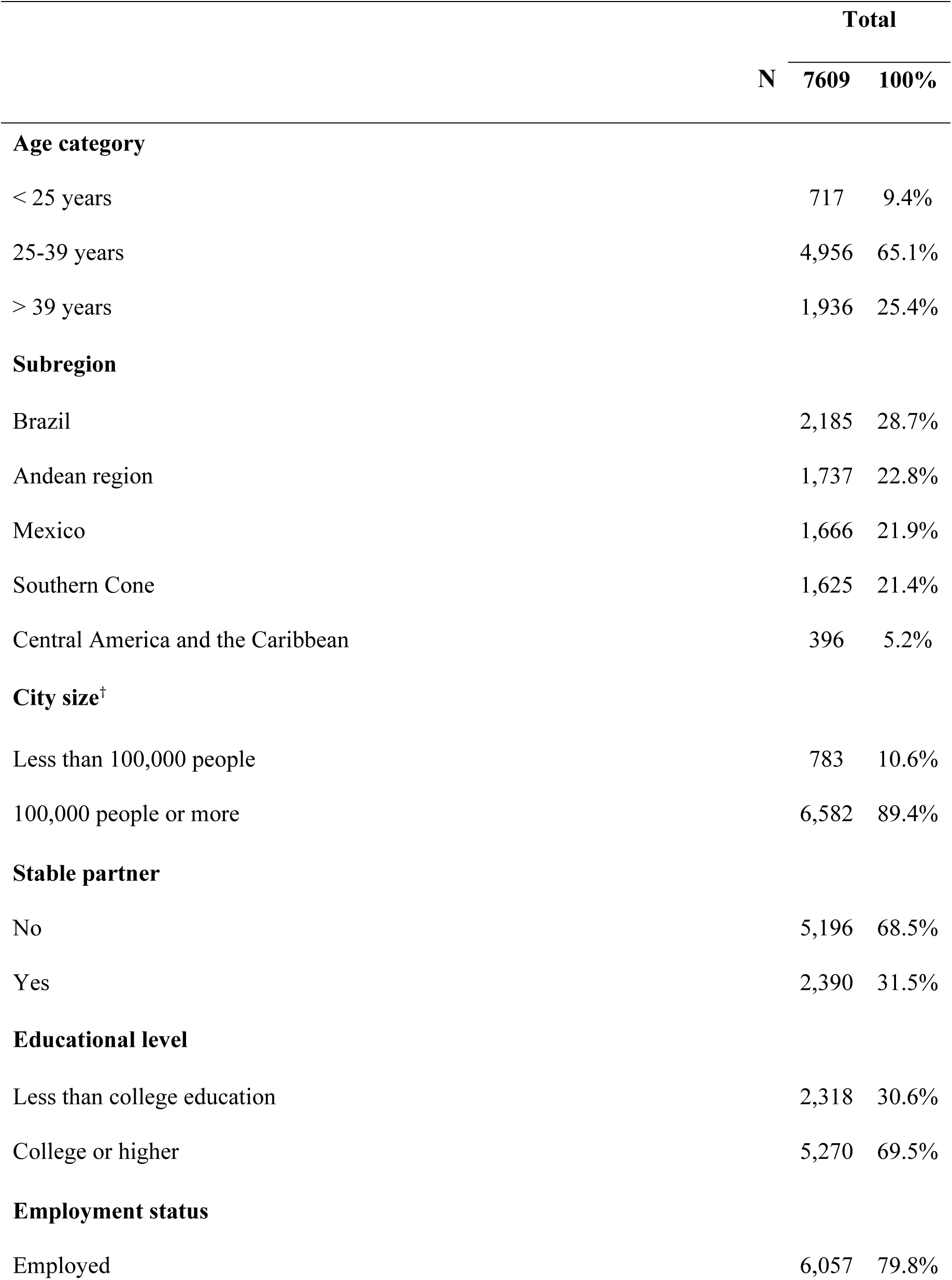
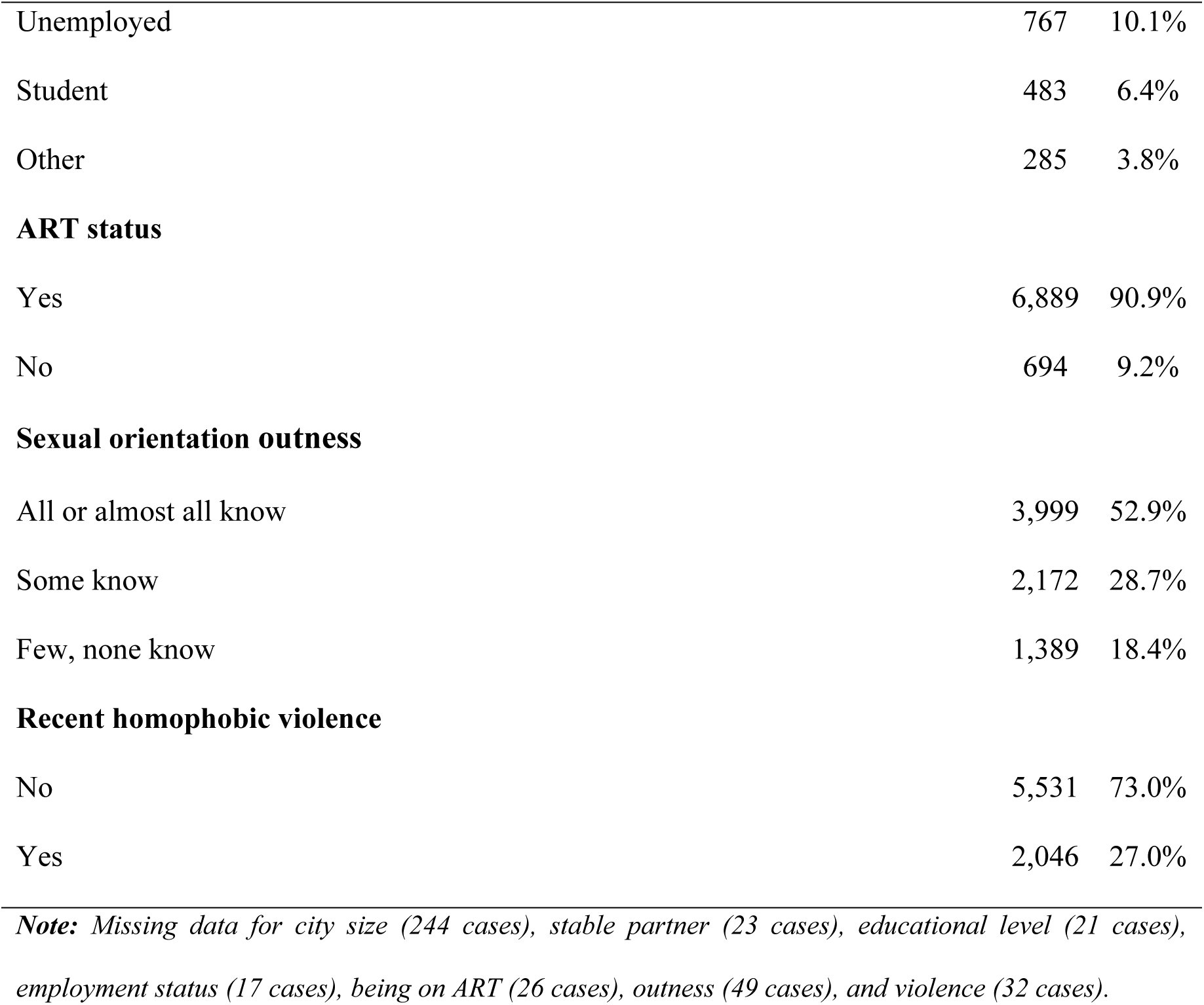
Participants’ characteristics.

|  |  | Total |  |
| --- | --- | --- | --- |
|  | N | 7609 | 100% |
| Age category |  |  |  |
| < 25 years |  | 717 | 9.4% |
| 25-39 years |  | 4,956 | 65.1% |
| > 39 years |  | 1,936 | 25.4% |
| Subregion |  |  |  |
| Brazil |  | 2,185 | 28.7% |
| Andean region |  | 1,737 | 22.8% |
| Mexico |  | 1,666 | 21.9% |
| Southern Cone |  | 1,625 | 21.4% |
| Central America and the Caribbean |  | 396 | 5.2% |
| City size<sup>†</sup> |  |  |  |
| Less than 100,000 people |  | 783 | 10.6% |
| 100,000 people or more |  | 6,582 | 89.4% |
| Stable partner |  |  |  |
| No |  | 5,196 | 68.5% |
| Yes |  | 2,390 | 31.5% |
| Educational level |  |  |  |
| Less than college education |  | 2,318 | 30.6% |
| College or higher |  | 5,270 | 69.5% |
| Employment status |  |  |  |
| Employed |  | 6,057 | 79.8% |

|  |  |  |
| --- | --- | --- |
| Unemployed | 767 | 10.1% |
| Student | 483 | 6.4% |
| Other | 285 | 3.8% |
| **ART status** |  |  |
| Yes | 6,889 | 90.9% |
| No | 694 | 9.2% |
| **Sexual orientation outness** |  |  |
| All or almost all know | 3,999 | 52.9% |
| Some know | 2,172 | 28.7% |
| Few, none know | 1,389 | 18.4% |
| **Recent homophobic violence** |  |  |
| No | 5,531 | 73.0% |
| Yes | 2,046 | 27.0% |
| ***Note:** Missing data for city size (244 cases), stable partner (23 cases), educational level (21 cases), employment status (17 cases), being on ART (26 cases), outness (49 cases), and violence (32 cases).* |  |  |

### Distribution of the Dependent and Independent Variables

The key sociodemographic characteristics of participants are presented by outcome and independent variables (Table 2). Regarding ART status, 15% of individuals under 25 years of age reported not being on ART, as did 9% of those aged 25 to 39 and 6% of those over 39 years. In the Andean countries, 13% of participants were not on ART, while in the Southern Cone the figure was 10%. Other regions reported proportions below 8%. Among participants living in cities with fewer than 100,000 inhabitants, 13% reported not being on ART. This was also observed among single participants (10%), those with less than a university education (12%), unemployed individuals (13%), and students (12%).

**Table 2.**
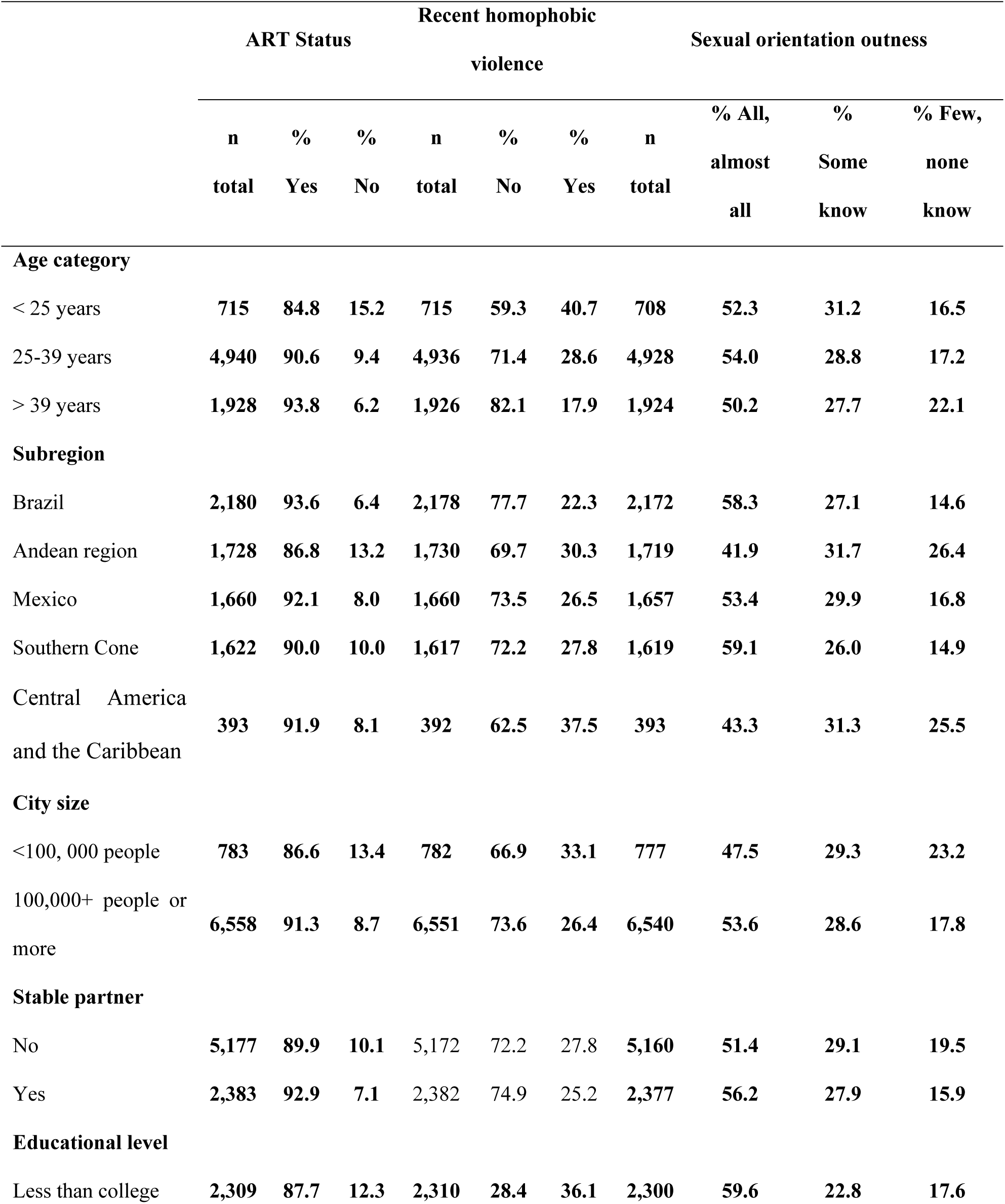
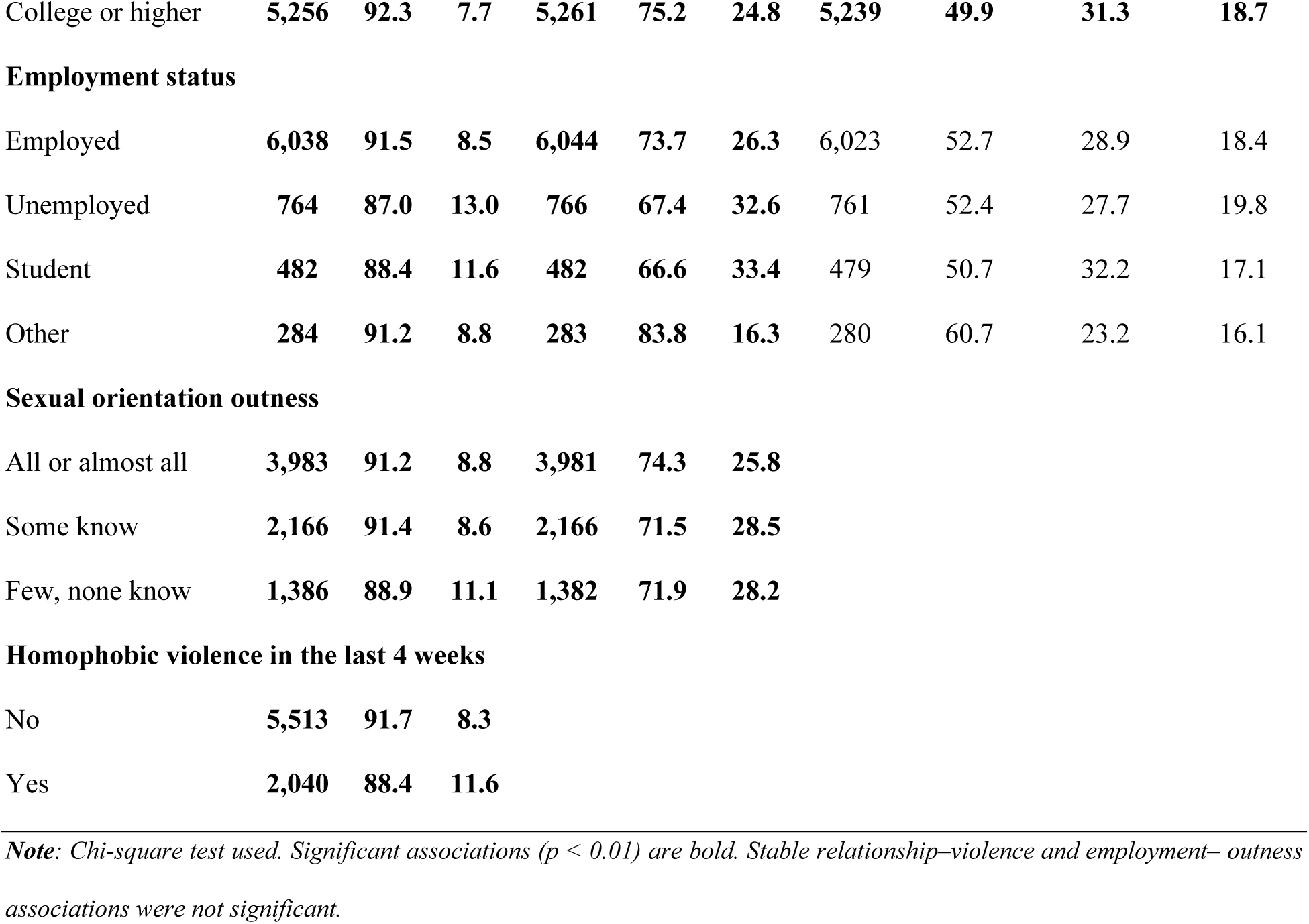
Participants’ Characteristics according to Not being on ART, Recent homophobic violence, and Sexual orientation outness.

|  | ART Status |  |  | Recent homophobic violence |  |  | Sexual orientation outness |  |  |  |
| --- | --- | --- | --- | --- | --- | --- | --- | --- | --- | --- |
|  | n | % | % | n | % | % | n | % All, | % | % Few, |
|  | total | Yes | No | total | No | Yes | total | almost all | Some know | none know |
| **Age category** |  |  |  |  |  |  |  |  |  |  |
| < 25 years | 715 | 84.8 | 15.2 | 715 | 59.3 | 40.7 | 708 | 52.3 | 31.2 | 16.5 |
| 25-39 years | 4,940 | 90.6 | 9.4 | 4,936 | 71.4 | 28.6 | 4,928 | 54.0 | 28.8 | 17.2 |
| > 39 years | 1,928 | 93.8 | 6.2 | 1,926 | 82.1 | 17.9 | 1,924 | 50.2 | 27.7 | 22.1 |
| **Subregion** |  |  |  |  |  |  |  |  |  |  |
| Brazil | 2,180 | 93.6 | 6.4 | 2,178 | 77.7 | 22.3 | 2,172 | 58.3 | 27.1 | 14.6 |
| Andean region | 1,728 | 86.8 | 13.2 | 1,730 | 69.7 | 30.3 | 1,719 | 41.9 | 31.7 | 26.4 |
| Mexico | 1,660 | 92.1 | 8.0 | 1,660 | 73.5 | 26.5 | 1,657 | 53.4 | 29.9 | 16.8 |
| Southern Cone | 1,622 | 90.0 | 10.0 | 1,617 | 72.2 | 27.8 | 1,619 | 59.1 | 26.0 | 14.9 |
| Central America and the Caribbean | 393 | 91.9 | 8.1 | 392 | 62.5 | 37.5 | 393 | 43.3 | 31.3 | 25.5 |
| **City size** |  |  |  |  |  |  |  |  |  |  |
| <100, 000 people | 783 | 86.6 | 13.4 | 782 | 66.9 | 33.1 | 777 | 47.5 | 29.3 | 23.2 |
| 100,000+ people or more | 6,558 | 91.3 | 8.7 | 6,551 | 73.6 | 26.4 | 6,540 | 53.6 | 28.6 | 17.8 |
| **Stable partner** |  |  |  |  |  |  |  |  |  |  |
| No | 5,177 | 89.9 | 10.1 | 5,172 | 72.2 | 27.8 | 5,160 | 51.4 | 29.1 | 19.5 |
| Yes | 2,383 | 92.9 | 7.1 | 2,382 | 74.9 | 25.2 | 2,377 | 56.2 | 27.9 | 15.9 |
| **Educational level** |  |  |  |  |  |  |  |  |  |  |
| Less than college | 2,309 | 87.7 | 12.3 | 2,310 | 28.4 | 36.1 | 2,300 | 59.6 | 22.8 | 17.6 |
| College or higher | **5,256** | **92.3** | **7.7** | **5,261** | **75.2** | **24.8** | **5,239** | **49.9** | **31.3** | **18.7** |
| **Employment status** |  |  |  |  |  |  |  |  |  |  |
| Employed | **6,038** | **91.5** | **8.5** | **6,044** | **73.7** | **26.3** | 6,023 | 52.7 | 28.9 | 18.4 |
| Unemployed | **764** | **87.0** | **13.0** | **766** | **67.4** | **32.6** | 761 | 52.4 | 27.7 | 19.8 |
| Student | **482** | **88.4** | **11.6** | **482** | **66.6** | **33.4** | 479 | 50.7 | 32.2 | 17.1 |
| Other | **284** | **91.2** | **8.8** | **283** | **83.8** | **16.3** | 280 | 60.7 | 23.2 | 16.1 |

**Sexual orientation outness**
|  |  |  |  |  |  |  |
| --- | --- | --- | --- | --- | --- | --- |
| All or almost all | **3,983** | **91.2** | **8.8** | **3,981** | **74.3** | **25.8** |
| Some know | **2,166** | **91.4** | **8.6** | **2,166** | **71.5** | **28.5** |
| Few, none know | **1,386** | **88.9** | **11.1** | **1,382** | **71.9** | **28.2** |

**Homophobic violence in the last 4 weeks**
|  |  |  |  |
| --- | --- | --- | --- |
| No | **5,513** | **91.7** | **8.3** |
| Yes | **2,040** | **88.4** | **11.6** |

*Note: Chi-square test used. Significant associations ( $p < 0.01$ ) are bold. Stable relationship–violence and employment– outness associations were not significant.*

Experiences of homophobic violence in the last 4 weeks, were reported by 40% of participants under 25 years of age; 30% of residents in the Andean countries, and 37.5% of residents in Central America and the Caribbean, 33% of those living in cities with fewer than 100,000 inhabitants, 36% of those without a university education, 28% of single participants, and 33% of both unemployed individuals and students. All associations were statistically significant (*p<0.01*), except that with stable partnership status, which was not significant.

Regarding the sexual orientation outness, 22% of individuals over 39 years of age; and 17% of both those aged 25-39 and those 24 or younger reported that few or no people knew about their orientation. In the Andean countries and Central America and the Caribbean, 26% of participants reported that few or no one knew about their orientation. Among those living in cities with fewer than 100,000 inhabitants, 23% reported low outness, as did 20% of single participants. Among individuals with higher education, 60% reported that most or all people knew about their orientation. Excluding employment status, all associations were statistically significant (*p<0.01*).

Those who experienced recent homophobic violence reported more frequently not being on ART (12% vs. 8%); likewise, those who reported that few/no one knew about their orientation, compared to the two other groups, reported more often not being on ART (11% vs. 9%). Additionally, 28-29% of participants who indicated that some or few/no one knew about their orientation reported experiencing rapid homophobic violence.

### Crude and adjusted prevalence ratio analysis

The results of the unadjusted PR models examining the association between ART status and other variables are presented in Table 3. All were statistically significant (p<0.001). Participants who reported that few or no one knew about their sexual orientation showed a higher prevalence of not being on ART (PR=1.26, 95% CI [1.06-1.51]), compared to those who reported that most or all knew. Those who reported homophobic violence in the last 4 weeks had a higher prevalence or not being on ART (PR=1.40, 95% CI [1.21-1.63]), compared to those who did not.

**Table 3.** Assessment of bivariable and multivariable relationships between Not being on ART, independent variables, and key sociodemographic factors.

| Exposures | Crude PR |  |  | Adjusted PR |  |  |
| --- | --- | --- | --- | --- | --- | --- |
|  | PR | 95% CI |  | PR | 95% CI |  |
|  |  | Lower | Upper |  | Lower | Upper |
|  |  | limit | limit |  | limit | limit |
| Sexual orientation outness |  |  |  |  |  |  |
| All or almost all | 1 |  |  | 1 |  |  |
| Some know | 0.98 | 0.82 | 1.16 | 0.98 | 0.82 | 1.17 |
| Few, none know | 1.26** | 1.06 | 1.51 | 1.22* | 1.01 | 1.48 |
| Recent homophobic violence |  |  |  |  |  |  |
| No | 1 |  |  | 1 |  |  |
| Yes | 1.40** | 1.21 | 1.63 | 1.25** | 1.07 | 1.46 |
| Age category |  |  |  |  |  |  |
| < 25 years | 1 |  |  | 1 |  |  |
| 25-39 years | 0.62*** | 0.51 | 0.75 | 0.71** | 0.58 | 0.89 |
| > 39 years | 0.41*** | 0.32 | 0.52 | 0.50*** | 0.38 | 0.65 |
| City size |  |  |  |  |  |  |
| Less than 100,000 people | 1 |  |  | 1 |  |  |
| 100,000 people or more | 0.65*** | 0.53 | 0.79 | 0.72*** | 0.59 | 0.87 |
| Stable partner |  |  |  |  |  |  |
| No | 1 |  |  | 1 |  |  |
| Yes | 0.71*** | 0.60 | 0.84 | 0.78** | 0.66 | 0.93 |
| Educational level |  |  |  |  |  |  |
| Less than college | 1 |  |  | 1 |  |  |
| College or higher | 0.63*** | 0.54 | 0.72 | 0.74*** | 0.63 | 0.86 |
| **Employment Status** |  |  |  |  |  |  |
| Employed | 1 |  |  | 1 |  |  |
| Unemployed | 1.53*** | 1.25 | 1.87 | 1.26* | 1.02 | 1.56 |
| Student | 1.37* | 1.05 | 1.77 | 0.97 | 0.73 | 1.29 |
| Other | 1.04 | 0.71 | 1.52 | 1.30 | 0.87 | 1.92 |
| **Subregion** |  |  |  |  |  |  |
| Brazil | 1 |  |  | 1 |  |  |
| Andean Region | 2.05*** | 1.68 | 2.51 | 1.87*** | 1.50 | 2.32 |
| Mexico | 1.24 | 0.98 | 1.56 | 1.28* | 1.01 | 1.62 |
| Southern Cone | 1.56*** | 1.25 | 1.93 | 1.45*** | 1.16 | 1.82 |
| Central America and the Caribbean | 1.27 | 0.88 | 1.83 | 0.99 | 0.66 | 1.47 |

\* $p < 0.05$ , \*\* $p < 0.01$ , \*\*\* $p < 0.001$

Other factors associated with not being on ART included unemployment (PR =1.53, 95% CI [1.25-1.87]) and being a student (PR =1.37, 95% CI [1.05-1.77], p=0.018) compared to being an employee. Participants from the Andean countries (PR =2.05, 95% CI [1.68-2.51]) and the Southern Cone (PR =1.56, 95% CI [1.25-1.93]) were more likely to not being on ART, compared to those from Brazil.

Participants aged 25-39 years (PR =0.62, 95% CI [0.51-0.75]) and those aged 39 years or older (PR =0.41, 95% CI [0.32-0.52]) had a lower prevalence of not being on ART compared to those under 25 years old. Those living in cities with more than 100,000 people (PR =0.62, 95% CI [0.51-0.75]), those in a relationship (PR =0.71, 95% CI [0.60-0.84]), and those with higher education (PR =0.63, 95% CI [0.54-0.72]) had also a lower prevalence of not being on ART.

The results of the multivariable analysis are presented in Table 3, corresponding to the first model, in which subregions were included in the analysis. Regarding the primary outcome, both reporting that few or no one knew about their sexual orientation (aPR =1.22, 95% CI [1.01-1.48], p=0.043) and having experienced homophobic violence in the past 4 weeks (aPR=1.25, 95% CI [1.07-1.46], p=0.005) were associated with a higher prevalence of not being on ART. Significant aPRs were also observed for other characteristics, with inverse associations for factors such as being aged 25–39 (aPR=0.71, 95% CI [0.58-0.89], p=0.002) or over 39 years old (aPR=0.50, 95% CI [0.38-0.65], p<0.001) compared to those under 25, living in a city with more than 100,000 people (aPR=0.72, 95% CI [0.59-0.87], p=0.001), having a stable partner (aPR=0.78, 95% CI [0.66-0.93], p=0.005), and having a university education (aPR=0.74, 95% CI [0.63-0.86], p<0.001). Direct associations were found with being unemployed (aPR=1.26, 95% CI [1.02-1.56], p=0.030) and living in countries within the Andean subregion (aPR=1.89, 95% CI [1.52-2.34], p<0.001), Mexico (aPR=1.28, 95% CI [1.01-1.62], p=0.040), and the Southern Cone (aPR=1.45, 95% CI [1.16-1.82], p=0.001).

Another important aspect to highlight is the proportion of participants not being on ART and the aPRs by country, estimated after controlling for all variables included in the model. Although results have been presented by subregions, examining country-level findings is relevant as a proxy for differences in the performance and capacity of national health systems. Table 4 presents the results of a second multivariable model, in which countries, rather than subregions, were included in the analysis. In this table, substantial heterogeneity is observed across countries, with significantly higher prevalence of not being on ART in Venezuela (aPR=3.20, 95% CI [2.43–4.21], p=0.001) and Paraguay (aPR=2.99, 95% CI [1.85–4.82], p=0.001), compared to Brazil. Other countries, such as Colombia (aPR=1.70, 95% CI [1.32–2.19], p=0.001) and Ecuador (aPR=1.79, 95% CI [1.15–2.78], p=0.01), also show higher likelihoods of not being on treatment, whereas results from Central American countries are not statistically significant. Although Suriname shows the highest adjusted prevalence ratio (aPR=6.32, 95% CI [2.87–13.94], p<0.001), this finding should be interpreted with caution due to the small sample size. Overall, these findings suggest important between-country inequalities, which may reflect structural differences in access to care, continuity of treatment, and the capacity of health systems to respond to HIV. In this second model, the aPRs of the other variables remained at similar levels of statistical significance, with no substantial changes in confidence intervals or point estimates compared to the first model.

**Table 4.** Assessment of multivariable relationships between Not being on ART by country.

| Exposures |  | ART Status |  | Adjusted PR |  |  |
| --- | --- | --- | --- | --- | --- | --- |
|  |  |  |  | 95% CI |  |  |
| Subregion | Pais | n total | % No | PR | Lower limit | Upper limit |
| *Brazil* | Brazil | 2,180 | 6.4 | 1 |  |  |
| *Andean Region* | Bolivia | 57 | 14.0 | 1.80 | 0.93 | 3.50 |
|  | **Ecuador** | **143** | **15.4** | **1.79\*\*** | **1.15** | **2.78** |
|  | Perú | 289 | 8.0 | 1.03 | 0.66 | 1.62 |
|  | **Colombia** | **910** | **11.4** | **1.70\*\*\*** | **1.32** | **2.19** |
|  | **Venezuela** | **329** | **21.6** | **3.20\*\*\*** | **2.43** | **4.21** |
| *Mexico* | **Mexico** | **1,660** | **8.0** | **1.28\*** | **1.01** | **1.63** |
| *Southern Cone* | **Argentina** | **750** | **9.7** | **1.38\*** | **1.03** | **1.84** |
|  | **Chile** | **719** | **8.9** | **1.36\*** | **1.02** | **1.82** |
|  | **Paraguay** | **78** | **20.5** | **2.99\*\*\*** | **1.85** | **4.82** |
|  | Uruguay | 75 | 12.0 | 1.45 | 0.71 | 2.96 |
| *Central America and the Caribbean* | Costa Rica | 102 | 10.8 | 1.12 | 0.56 | 2.22 |
|  | El Salvador | 46 | 8.7 | 1.16 | 0.45 | 2.98 |
|  | Guatemala | 98 | 3.1 | 0.46 | 0.15 | 1.42 |
|  | Honduras | 35 | 11.4 | 1.58 | 0.63 | 3.98 |
|  | Nicaragua | 15 | 0.0 | - | - | - |
|  | Panama | 85 | 5.9 | 0.68 | 0.25 | 1.79 |
|  | Suriname | **12** | **41.7** | **6.32\*\*\*** | **2.87** | **13.94** |

\* $p<0.05$ , \*\* $p<0.01$ , \*\*\* $p<0.001$

## Discussion

We found that 9% of MSM diagnosed with HIV at least one year prior to data collection in the region were not on ART—an estimate that may hinder progress toward achieving the 95% treatment target set by UNAIDS, highlighting the limited progress in recent years[17]. This situation could be partly explained by the persistent underfunding of HIV services for key populations and the lack of detailed strategic information on these groups, factors highlighted by UNAIDS in recent years[6].

MSM who reported recent homophobic violence or low sexual orientation outness were less likely to be on ART. Other directly associated factors identified included unemployment, residing in Mexico or countries in the Andean subregion (Ecuador, Colombia and Venezuela) or the Southern Cone (Argentina, Chile and Paraguay). Inverse associations were found by reporting higher educational attainment, having a stable partner, living in large cities, and being over 25 years of age.

These findings are consistent with conceptual frameworks suggesting that intersecting forms of violence may shape HIV care outcomes through psychosocial and structural pathways, including mental health, internalized stigma, and reduced trust in health systems[32]. In this context, homophobic violence and limited sexual orientation outness may represent mechanisms influencing disengagement from ART among MSM in LA.

These findings are consistent with studies from other regions showing that discrimination[13,26,28,35], low sexual orientation outness or concealment of HIV status[28–30,46], and experiences of violence related to disclosure[35] negatively impact adherence and retention in care. Regarding sociodemographic variables, factors most strongly associated with not being on ART reported previously include being male[16,24,30,47–50], younger age[13,15,16,22,24,29,30,34,48,50,51], lower educational attainment[12,52,53], being unemployed[12,15,22,24,53], lacking financial support[15,26], not having a social support network such as a partner[24,26,49,52], being an immigrant[15,53], and lacking a stable residence[53]. Living far from healthcare services where ART is provided is another identified risk factor[24,26,30,47,48,50–55].

Recent experiences of homophobic violence may reflect ongoing victimization and serve as a proxy for current exposure to structural homophobia. Discrimination and violence based on sexual orientation can limit access to and use of HIV prevention, testing, and treatment services[56,57]. In such contexts, MSM may face barriers related to the risk of involuntary disclosure of their sexual orientation, with potential social, familial, and professional consequences. Greater outness may also increase exposure to homophobic violence, particularly among younger individuals, who may have fewer resources, greater dependence on family or social environments, and reduced capacity to cope with or avoid such situations. As a result, some individuals may have low sexual orientation outness or avoid healthcare settings where they could be recognized, including HIV prevention and treatment services. This may be more pronounced in smaller cities or rural areas, where the risk of being recognized may be higher and services may be less available, particularly for younger individuals, potentially reducing service utilization and negatively affecting treatment initiation and continuity. Although regional indicators such as the LGBT Equality Index suggest relatively higher levels of protection in parts of LA compared to other regions[58], important within-region disparities persist, particularly in some Andean and Central American countries.

Higher education, living in urban areas, and having a social support network may provide individuals with resources that facilitate access to healthcare. These factors are likely associated with greater exposure to treatment-related information, including the benefits of sustained adherence such as Undetectable = Untransmissible. Economic resources and proximity to services in densely populated areas may further facilitate healthcare access, while emotional support from family or a partner can support ART initiation and continuity. Lower levels of health investment in Andean countries[59,60] and less inclusive social environments for MSM[58] may contribute to challenges in retaining and re-engaging MSM in HIV care. Although countries in the Southern Cone and Mexico show a greater presence of community-based sexual health organizations[61] and more inclusive policies toward MSM[58], residing in these countries was still associated with a higher prevalence of not being on ART, compared to Brazil. Therefore, understanding both loss to follow-up and disruptions in ART continuity requires attention to dynamic sociocultural processes shaped negatively by stigma, discrimination, and violence, and positively by protective factors such as social support, particularly among men[62].

Although the performance of national health systems was not directly measured, the country-level analysis provides a proxy for their influence on access to ART. Substantial heterogeneity was observed, with higher prevalence of not being on treatment in Andean countries (Venezuela, Colombia, and Ecuador), Southern Cone countries (Paraguay, Argentina, and Chile), and Mexico. In Venezuela, this may reflect health system deterioration associated with interruptions in ART availability[63]. In other countries, despite free ART provision, gaps in effective access and timely treatment initiation persist like in Colombia or Ecuador[64]. In Chile and Argentina, where coverage is high, this pattern may instead reflect challenges in retention and adherence to treatment[65].

This regional study has some limitations. The cross-sectional design restricts causal inference to hypothesis generation. The use of an online survey resulted in a convenience sample that excluded individuals without digital access, and the study was not designed to ensure representativeness of people living with HIV, thereby limiting generalizability. Recruitment through dating applications may have facilitated participation among individuals less open about their sexual orientation, while excluding those with lower sexual activity and potentially different health-related behaviors. Given inter-country variability in sociocultural determinants and health systems (for example, the impact of political crisis on Venezuela health system, etc.), associations with macro-region should be taken with caution, as these differences may influence the observed results and their interpretation beyond individual-level factors; further analysis will be needed to have a better understanding of our findings at a country level. Rurality was not directly measured; instead, city size (100,000-inhabitant threshold) was used as a proxy, which may not fully capture heterogeneity in access to HIV services across settings.

The LAMIS-2018 study provided a unique opportunity to investigate HIV in LA, as it represents the first regional survey allowing the evaluation of underexplored, crucial topics for one of the key populations most affected by HIV, using a common instrument. Among the most relevant topics explored were the initiation and discontinuation of antiretroviral treatment among MSM living with HIV, as well as discrimination, homophobic violence, and sexual orientation outness, all of which are key but underexplored areas in the region.

### Conclusions

Findings from this study indicate that recent homophobic violence and low sexual orientation outness were associated with not being on ART among MSM living with HIV in Latin America. While access to ART may vary across countries according to health system characteristics, these results suggest that structural factors such as homophobia, stigma, and violence may limit access to ART. In this context, not being on ART may reflect the combined influence of health system differences and these social processes. Addressing these challenges may require multisectoral approaches that strengthen health systems while also reducing stigma and violence to improve access to and continuity of HIV care among key populations, which will be critical to advancing progress toward the 95–95–95 targets.

## Data Availability

This study did not generate primary data. Secondary data from the LAMIS-2018 database were used, which can be requested from the study coordinators through the following form: https://docs.google.com/document/d/1gHoX_E90jyrMTokzSJWT6OfZs3_0icB_/edit.”

## Competing interest’s

The authors have no competing interests to declare

## Authors’ contributions

JE conceived and designed the study, performed the data analysis, and wrote the manuscript as part of his master’s thesis. AS was a co-advisor for the study, while ER provided advice on the statistical aspects. LH participated as a reviewer and advisor in the manuscript writing process. CC was the main advisor for the study. All authors reviewed and approved the manuscript before submission.

## Author information

JE conducted the study as part of his thesis for the Master’s degree in Control of Infectious and Tropical Diseases (MCEIT) at UPCH. JE, CC and ER participated in the LAMIS-2018 parent study as investigators from Peru. CC was one of the principal investigators of the parent study, and ER oversaw the statistical analysis and writing of the LAMIS-2018 regional report.

## Acknowledgements

We would like to thank the RIGHT PLUS study team of Coalition PLUS for granting access and approving the use of the LAMIS-2018 database for this manuscript. We would also like to thank MCEIT for providing advice on conducting the study. We gratefully acknowledge the support of the Vice-Rectorate of Research at Universidad Peruana Cayetano Heredia for their assistance in the design and development of this scientific article. Finally, we thank all the sexual diversity community organizations in the 18 countries that supported the dissemination and promotion of the survey.

## Funding

This study was carried out as part of a MCEIT program. The main author was able to carry out this program thanks to funding from the PARACAS Scholarship (Program for Advanced Research Capacities for AIDS in Peru) and the Interdisciplinary Research Center on Sexuality, AIDS and Society – UPCH.

## Disclaimer

None

## Data Availability Statement

The database used for this study is available upon request from interested parties at: https://www.coalitionplus.org/lamis/

## Supporting Information

Supporting Information file 1: Stata codes of the study

Information on file format. Do.file file in STATA that contains all the codes used to carry out this study.

## List of abbreviations

ART: Antiretroviral Therapy
aPR: Adjusted Prevalence Ratios
CAGE-4: Cut down, Annoyed, Guilty, Eye-opener
HIV: Human Immunodeficiency Virus
LA: Latin America
LAMIS: Latin American MSM Internet Survey
LGBTQ+: Lesbian, Gay, Bisexual, Transgender, Queer and more
MSM: Men who have sex with men
PHQ-4: Patient Health Questionnaire-4
PLHIV: People living with HIV
PR: Prevalence Ratios
RIGHT PLUS: Ibero-American Network of Studies on Gay Men, Other Men Who Have Sex with Men, and Transgender People
’U=U’: Undetectable = Untransmittable
UNAIDS: Joint United Nations Programme on HIV/AIDS

## Notes

### Competing Interest Statement

The authors have declared no competing interest.

### Funding Statement

The author(s) received no specific funding for this work.

### Author Declarations

The Institutional Ethics Committee of Universidad Peruana Cayetano Heredia (code: CIEI-196-20-24).

## References

1. Crabtree-Ramírez B, Belaunzarán-Zamudio PF, Cortes CP, Morales M, Sued O, Sierra-Madero J, et al. The HIV epidemic in Latin America: a time to reflect on the history of success and the challenges ahead. J Int AIDS Soc. 2020;23:e25468. 10.1002/jia2.25468

2. UNAIDS. FACT SHEET 2024 [Internet]. 2024. https://www.unaids.org/sites/default/files/media_asset/UNAIDS_FactSheet_en.pdf

3. UNAIDS. UNAIDS DATA 2022 [Internet]. Geneva: Joint United Nations Programme on HIV/AIDS; 2022. https://www.unaids.org/sites/default/files/media_asset/data-book-2022_en.pdf

4. ONUSIDA. APROVECHANDO EL MOMENTO: La respuesta al VIH en América Latina [Internet]. 2020. https://www.unaids.org/sites/default/files/media_asset/2020_global-aids-report-latin-america_es.pdf

5. UNAIDS. The urgency of now: AIDS at a crossroads [Internet]. Geneva; 2024. https://www.unaids.org/sites/default/files/media_asset/2024-unaids-global-aids-update_en.pdf

6. UNAIDS. CONFRONTING INEQUALITIES: Lessons for pandemic responses from 40 years of AIDS [Internet]. 20 Avenue Appia 1211 Geneva 27 Switzerland: Joint United Nations Programme on HIV/AIDS; 2021 p. 386. https://www.unaids.org/sites/default/files/media_asset/2021-global-aids-update_en.pdf

7. Link B, Phelan J. Stigma and Its Public Health Implications. Lancet. 2006;367:528–9. 10.1016/S0140-6736(06)68184-1

8. UNAIDS. HIV AND STIGMA AND DISCRIMINATION [Internet]. 2024. https://www.unaids.https://www.unaids.org/sites/default/files/media_asset/07-hiv-human-rights-factsheet-stigma-discrmination_en.pdf

9. UNAIDS. Understanding Fast-Track: Accelerating Action to End the AIDS Epidemic by 2030 [Internet]. 20 Avenue Appia 1211 Geneva 27 Switzerland: Joint United Nations Programme on HIV/AIDS; 2015 p. 10. https://www.unaids.org/sites/default/files/media_asset/201506_JC2743_Understanding_FastTrack_en.pdf

10. UNAIDS. Global AIDS Strategy 2021-2026 — End Inequalities. End AIDS. [Internet]. 20 Avenue Appia 1211 Geneva 27 Switzerland: Joint United Nations Programme on HIV/AIDS; 2021 p. 160. https://www.unaids.org/sites/default/files/media_asset/global-AIDS-strategy-2021-2026_en.pdf

11. Bazán-Ruiz S, Chanamé Pinedo LE, Maguiña Vargas C. Adherencia al TARGA en VIH /SIDA. Un Problema de Salud Pública. Acta Med Per. 2013;30:102–3.

12. Ibiloye O, Decroo T, Eyona N, Eze P, Agada P. Characteristics and early clinical outcomes of key populations attending comprehensive community-based HIV care: Experiences from Nasarawa State, Nigeria. Mugo PM, editor. PLoS ONE. 2018;13:e0209477. 10.1371/journal.pone.0209477

13. Ajeh RA, Gregory HE, Thomas EO, Noela NA, Dzudie A, Jules AN, et al. Determinants of retention in HIV antiretroviral treatment (ART) in the Cameroon International epidemiology Database to Evaluate AIDS (IeDEA) study clinics: the context of the HIV treat all strategy in Cameroon. Pan Afr Med J. 2021;40:129. 10.11604/pamj.2021.40.129.22642

14. Vivancos MJ, Moreno S. Follow-up losses of people with HIV infection: A weak point in the continuum of care. Enferm Infecc Microbiol Clin (Engl Ed). 2019;37:359–60. 10.1016/j.eimc.2019.03.006

15. Teira R, Espinosa N, Gutiérrez MM, Montero M, Martínez E, González F, et al. Losses to follow-up of HIV-infected people in the Spanish VACH cohort over the period between 2013 and 2014: The importance of sociodemographic factors. Enferm Infecc Microbiol Clin (Engl Ed). 2019;37:361–6. 10.1016/j.eimc.2018.09.008

16. Inzaule SC, Kroeze S, Kityo CM, Siwale M, Akanmu S, Wellington M, et al. Long-term HIV treatment outcomes and associated factors in sub-Saharan Africa: multicountry longitudinal cohort analysis. AIDS. 2022;36:1437–47. 10.1097/QAD.0000000000003270

17. Fox MP, Rosen S. Retention of Adult Patients on Antiretroviral Therapy in Low- and Middle-Income Countries: Systematic Review and Meta-analysis 2008-2013. J Acquir Immune Defic Syndr. 2015;69:98–108. 10.1097/QAI.0000000000000553

18. World Health Organization. Global health sector strategy on HIV 2016–2021: towards ending AIDS [Internet]. Geneva; 2016 may. https://www.who.int/publications/i/item/WHO-HIV-2016.05

19. Gallego N, Diaz A, Folch C, Meyer S, Vazquez M, Casabona J, et al. Factors associated with low levels of HIV testing among young men who have sex with men (MSM) participating in EMIS-2017 in Spain. Sex Transm Infect. 2022;98:518–24. 10.1136/sextrans-2021-055193

20. Millett WLJI Stephen A Flores, Cherie R Rooks-Peck, Deborah J Gelaude, Lisa Belcher, Philip M Ricks, Gregorio A. Experienced Homophobia and HIV Infection Risk Among U.S. Gay, Bisexual, and Other Men Who Have Sex with Men: A Meta-Analysis - William L. Jeffries, Stephen A. Flores, Cherie R. Rooks-Peck, Deborah J. Gelaude, Lisa Belcher, Philip M. Ricks, Gregorio A. Millett, 2021. LGBT Health [Internet]. 2021 [citado 15 de abril de 2026]; https://journals.sagepub.com/doi/10.1089/lgbt.2020.0274. Accedido 15 abr 2026

21. Tatés-Ortega N, Álvarez J, López L, Mendoza-Ticona A, Alarcón-Arrascue E. Pérdida en el seguimiento de pacientes tratados por tuberculosis resistente a rifampicina o multidrogorresistente en Ecuador. Rev Panam Salud Publica. 2019;43:e91. 10.26633/RPSP.2019.91

22. Támara-Ramírez JR, Álvarez CA, Rodríguez J. Pérdida de seguimiento y factores asociados en pacientes inscritos en el programa de HIV/sida del Hospital Universitario San Ignacio, Colombia, 2012-2013. biomedica. 2016;36:265. 10.7705/biomedica.v36i2.2939

23. De Boni RB, Peratikos MB, Shepherd BE, Grinsztejn B, Cortés C, Padgett D, et al. Is substance use associated with HIV cascade outcomes in Latin America? PLoS One. 2018;13:e0194228. 10.1371/journal.pone.0194228

24. Kebede HK, Mwanri L, Ward P, Gesesew HA. Predictors of lost to follow up from antiretroviral therapy among adults in sub-Saharan Africa: a systematic review and meta-analysis. Infect Dis Poverty. 2021;10:33. 10.1186/s40249-021-00822-7

25. Williams PL, Chernoff M, Angelidou K, Brouwers P, Kacanek D, Deygoo NS, et al. Participation and retention of youth with perinatal HIV infection in mental health research studies: the IMPAACT P1055 psychiatric comorbidity study. J Acquir Immune Defic Syndr. 2013;63:401–9. 10.1097/QAI.0b013e318293ad53

26. Adelekan B, Andrew N, Nta I, Gomwalk A, Ndembi N, Mensah C, et al. Social barriers in accessing care by clients who returned to HIV care after transient loss to follow-up. AIDS Res Ther. 2019;16:17. 10.1186/s12981-019-0231-5

27. Okonji EF, Mukumbang FC, Orth Z, Vickerman-Delport SA, Van Wyk B. Psychosocial support interventions for improved adherence and retention in ART care for young people living with HIV (10–24 years): a scoping review. BMC Public Health. 2020;20:1841. 10.1186/s12889-020-09717-y

28. Wolf HT, Halpern-Felsher BL, Bukusi EA, Agot KE, Cohen CR, Auerswald CL. «It is all about the fear of being discriminated [against]…the person suffering from HIV will not be accepted»: a qualitative study exploring the reasons for loss to follow-up among HIV-positive youth in Kisumu, Kenya. BMC Public Health. 2014;14:1154. 10.1186/1471-2458-14-1154

29. Evangeli M, Newell M-L, McGrath N. Factors associated with pre-ART loss-to-follow up in adults in rural KwaZulu-Natal, South Africa: a prospective cohort study. BMC Public Health. 2016;16:358. 10.1186/s12889-016-3025-x

30. Govindasamy D, Ford N, Kranzer K. Risk factors, barriers and facilitators for linkage to antiretroviral therapy care: a systematic review. AIDS. 2012;26:2059–67. 10.1097/QAD.0b013e3283578b9b

31. Gesesew HA, Tesfay Gebremedhin A, Demissie TD, Kerie MW, Sudhakar M, Mwanri L. Significant association between perceived HIV related stigma and late presentation for HIV/AIDS care in low and middle-income countries: A systematic review and meta-analysis. PLoS One. 2017;12:e0173928. 10.1371/journal.pone.0173928

32. Quinn KG, Spector A, Takahashi L, Voisin DR. Conceptualizing the Effects of Continuous Traumatic Violence on HIV Continuum of Care Outcomes for Young Black Men Who Have Sex with Men in the United States. AIDS Behav. 2021;25:758–72. 10.1007/s10461-020-03040-8

33. Carriquiry G, Fink V, Koethe JR, Giganti MJ, Jayathilake K, Blevins M, et al. Mortality and loss to follow-up among HIV-infected persons on long-term antiretroviral therapy in Latin America and the Caribbean. J Int AIDS Soc. 2015;18:20016. 10.7448/IAS.18.1.20016

34. de Almeida MC, de Jesus Pedroso N, do Socorro Lina van Keulen M, Jácome GPO, Fernandes GC, Yokoo EM, et al. Loss to Follow-Up in a Cohort of HIV-Infected Patients in a Regional Referral Outpatient Clinic in Brazil. AIDS Behav. 2014;18:2387–96. 10.1007/s10461-014-0812-1

35. Ortiz-Hernández L, Pérez-Salgado D, Miranda-Quezada IP, Staines-Orozco MG, Compean-Dardón MS. Experiences of homophobia and adherence to antiretroviral treatment (ART) in men who have sex with men (MSM). Saude soc. Faculdade de Saúde Pública, Universidade de São Paulo. Associação Paulista de Saúde Pública.; 2021;30:e200235.

36. Reyes-Diaz M, Folch C, Celly A, Stuardo V, Veras V, Schmidt A, et al. LAMIS-2018: Encuesta latinoamericana por internet en hombres que tienen sexo con hombres. Informe Regional. [Internet]. Right PLUS/Coalition PLUS; 2022. https://drive.google.com/file/d/13Hhrqhk3gkuzrdXw3VDYfDA78X1MRE6e/view?usp=sharing&usp=embed_facebook. Accedido 5 dic 2024

37. RIGHT PLUS - ES [Internet]. Coalition PLUS. [citado 5 de diciembre de 2024]. https://www.coalitionplus.org/rightplus/. Accedido 5 dic 2024

38. Reyes-Díaz M, Celly A, Folch C, Lorente N, Stuardo V, Veras MA, et al. Latin American Internet Survey for Men who have Sex with Men (LAMIS-2018): Design, methods and implementation. PLOS ONE. Public Library of Science; 2022;17:e0277518. 10.1371/journal.pone.0277518

39. King EB, Reilly C, Hebl M. The Best of Times, the Worst of Times: Exploring Dual Perspectives of “Coming Out” in the Workplace. Group & Organization Management. SAGE Publications Inc; 2008;33:566–601. 10.1177/1059601108321834

40. Marcus U, Jonas K, Berg R, Veras MA, Caceres CF, Casabona J, et al. Association of internalised homonegativity with partner notification after diagnosis of syphilis or gonorrhoea among men having sex with men in 49 countries across four continents. BMC Public Health. 2023;23:8. 10.1186/s12889-022-14891-2

41. Bränström R, Fellman D, Pachankis J. Structural Stigma and Sexual Minority Victimization Across 28 Countries: The Moderating Role of Gender, Gender Nonconformity, and Socioeconomic Status. J Interpers Violence. 2022;38:3563–85. 10.1177/08862605221108087

42. Napper LE, Fisher DG, Reynolds GL, Johnson ME. HIV Risk Behavior Self-Report Reliability at Different Recall Periods. AIDS Behav. 2010;14:152–61. 10.1007/s10461-009-9575-5

43. Kroenke K, Spitzer RL, Williams JBW, Löwe B. An Ultra-Brief Screening Scale for Anxiety and Depression: The PHQ–4. Psychosomatics. 2009;50:613–21. 10.1016/S0033-3182(09)70864-3

44. Ewing JA. Detecting Alcoholism: The CAGE Questionnaire. JAMA. 1984;252:1905–7. 10.1001/jama.1984.03350140051025

45. Cárdenas Castro JM. Potencia estadística y cálculo del tamaño del efecto en G*Power: complementos a las pruebas de significación estadística y su aplicación en psicología. Salud soc. 2014;5:210–44. 10.22199/S07187475.2014.0002.00006

46. Charurat ME, Emmanuel B, Akolo C, Keshinro B, Nowak RG, Kennedy S, et al. Uptake of Treatment as Prevention for HIV and Continuum of Care Among HIV-Positive Men Who Have Sex With Men in Nigeria. JAIDS Journal of Acquired Immune Deficiency Syndromes. 2015;68:S114–23. 10.1097/QAI.0000000000000439

47. Koech E, Stafford KA, Mutysia I, Katana A, Jumbe M, Awuor P, et al. Factors Associated with Loss to Follow-Up Among Patients Receiving HIV Treatment in Nairobi, Kenya. AIDS Research and Human Retroviruses. 2021;37:642–6. 10.1089/aid.2020.0292

48. Tweya H, Oboho IK, Gugsa ST, Phiri S, Rambiki E, Banda R, et al. Loss to follow-up before and after initiation of antiretroviral therapy in HIV facilities in Lilongwe, Malawi. Beck EJ, editor. PLoS ONE. 2018;13:e0188488. 10.1371/journal.pone.0188488

49. Wekesa P, McLigeyo A, Owuor K, Mwangi J, Ngugi E. Survival probability and factors associated with time to loss to follow-up and mortality among patients on antiretroviral treatment in central Kenya. BMC Infect Dis. 2022;22:522. 10.1186/s12879-022-07505-0

50. Hong SY, Winston A, Mutenda N, Hamunime N, Roy T, Wanke C, et al. Predictors of loss to follow-up from HIV antiretroviral therapy in Namibia. PLoS One. 2022;17:e0266438. 10.1371/journal.pone.0266438

51. Gebremichael MA, Gurara MK, Weldehawaryat HN, Mengesha MM, Berbada DA. Predictors of Loss to Follow-Up among HIV-Infected Adults after Initiation of the First-Line Antiretroviral Therapy at Arba Minch General Hospital, Southern Ethiopia: A 5-Year Retrospective Cohort Study. Cavalcanti Rolla V, editor. BioMed Research International. 2021;2021:1–12. 10.1155/2021/8659372

52. Hassan AS, Fielding KL, Thuo NM, Nabwera HM, Sanders EJ, Berkley JA. Early loss to follow-up of recently diagnosed HIV-infected adults from routine pre-ART care in a rural district hospital in Kenya: a cohort study. Trop Med Int Health. 2012;17:82–93. 10.1111/j.1365-3156.2011.02889.x

53. Diaz A, Ten A, Marcos H, Gutiérrez G, González-García J, Moreno S, et al. Determinantes de la asistencia irregular a consulta médica en pacientes con infección por el virus de la inmunodeficiencia humana: resultados de la Encuesta Hospitalaria de pacientes con el virus de la inmunodeficiencia humana, 2002-2012. Enferm Infecc Microbiol Clin. 2015;33. 10.1016/j.eimc.2014.07.009

54. Tomescu S, Crompton T, Adebayo J, Kinge CW, Akpan F, Rennick M, et al. Factors associated with an interruption in treatment of people living with HIV in USAID-supported states in Nigeria: a retrospective study from 2000–2020. BMC Public Health. 2021;21:2194. 10.1186/s12889-021-12264-9

55. Ibiloye O, Jwanle P, Masquillier C, Van Belle S, Jaachi E, Amoo O, et al. Long-term retention and predictors of attrition for key populations receiving antiretroviral treatment through community-based ART in Benue State Nigeria: A retrospective cohort study. Scheibe A, editor. PLoS ONE. 2021;16:e0260557. 10.1371/journal.pone.0260557

56. Organización Panamericana de la Salud. Proyecto para la Provisión de Atención Integral a los hombres gay y otros hombres que tienen sexo con hombres (HSH) en América Latina y el Caribe [Internet]. Washington, D.C: OPS; 2010. https://www.paho.org/sites/default/files/Blueprint%20MSM%20Final%20SPANISH.pdf

57. UNAIDS. HIV AND GAY MEN AND OTHER MEN WHO HAVE SEX WITH MEN [Internet]. 2024. https://www.unaids.org/sites/default/files/media_asset/03-hiv-human-rights-factsheet-gay-men_en.pdf

58. Equaldex. LGBT Rights by Country & Travel Guide [Internet]. Equaldex. 2025 [citado 5 de enero de 2025]. https://www.equaldex.com/. Accedido 5 ene 2025

59. Health spending as percent of GDP in Latin America [Internet]. TheGlobalEconomy.com. [citado 6 de julio de 2025]. https://www.theglobaleconomy.com/rankings/health_spending_as_percent_of_gdp/Latin-Am/?utm_source=chatgpt.com. Accedido 6 jul 2025

60. OECD, The World Bank. Panorama de la Salud: Latinoamérica y el Caribe 2020 [Internet]. OECD; 2020 [citado 5 de enero de 2025]. 10.1787/740f9640-es

61. Silva-Santisteban A, Eng S, de la Iglesia G, Falistocco C, Mazin R. HIV prevention among transgender women in Latin America: implementation, gaps and challenges. J Int AIDS Soc. 2016;19:20799. 10.7448/IAS.19.3.20799

62. Stuardo Ávila V, Manriquez Urbina JM, Fajreldin Chuaqui V, Belmar Prieto J, Valenzuela Santibáñez V. Model of socio-cultural dimensions involved in adherence to antiretroviral therapy for HIV/AIDS in public health care centers in Chile. AIDS Care. Taylor & Francis; 2016;28:1441–7. 10.1080/09540121.2016.1179252

63. Page KR, Doocy S, Reyna Ganteaume F, Castro JS, Spiegel P, Beyrer C. Venezuela’s public health crisis: a regional emergency. The Lancet. 2019;393:1254–60. 10.1016/S0140-6736(19)30344-7

64. González-Duran JA, Plaza RV, Luna L, Arbeláez MP, Deviaene M, Keynan Y, et al. Delayed HIV treatment, barriers in access to care and mortality in tuberculosis/HIV co-infected patients in Cali, Colombia. Colomb Med (Cali). 52:e2024875. 10.25100/cm.v52i3.4875

65. Costa J de M, Torres TS, Coelho LE, Luz PM. Adherence to antiretroviral therapy for HIV/AIDS in Latin America and the Caribbean: Systematic review and meta-analysis. J Int AIDS Soc. 2018;21:e25066. 10.1002/jia2.25066

